# COVID-19 or seasonal influenza? How to distinguish in people younger than 65 years old — — A retrospective observational cohort study comparing the 2009 pandemic influenza A H1N1 with 2022 SARS-CoV-2 Omicron BA.2 outbreaks in China

**DOI:** 10.1101/2023.02.28.23286466

**Authors:** Wen Zhong, Yisong Wu, Wenxiang Yue, Jiabin Fang, Baosong Xie, Nengluan Xu, Ming lin, Xiongpeng Zhu, Zhijun Su, Yusheng Chen, Hong Li, Hongru Li

## Abstract

**Objective:** This study attempted to explore the difference of clinical characteristics in H1N1 influenza infection and SARS-CoV-2 Omicron infection in people younger than 65 years old, in order to better identify the two diseases.

**Methods:** A total of 127 H1N1 influenza patients diagnosed from May 2009 to July 2009 and 3265 patients diagnosed and identified as SARS-CoV-2 Omicron BA.2 variant from March 2022 to May 2022 were admitted in this study. Through the 1 : 2 match based on age (The difference is less than 2 years), gender and underlying diseases, 115 patients with H1N1 infection and 230 patients with SARS-CoV-2 Omicron BA.2 infection(referred to as H1N1 group and Omicron group) were included in the statistics. The clinical manifestations of H1N1 group were compared with those of Omicron group. Logistic regression was performed to analyze the possible independent risk factors of H1N1 group and Omicron group. And multiple linear regression was used to analyze the factors for time for nucleic acid negativization (NAN).

**Results:** The median age of the two groups was 21 [11,26] years. Compared with the H1N1 group, the Omicron group had lower white blood cell count and CRP levels, less fever, nasal congestion, sore throat, cough, sputum and headache, while more olfactory loss, muscle soreness and LDH abnormalities. The Omicron group used less antibiotics and antiviral drugs, and the NAN time was longer (17 [14,20] VS 4 [3,5], P < 0.001). After logistic regression, it was found that fever, cough, headache, and increased white blood cell count were more correlated with the H1N1 group, while muscle soreness and LDH abnormalities were more correlated with the Omicron group. After analyzing the factors of NAN time, it was found that fever (B 1.529, 95 % CI [0.149,2.909], P = 0.030) significantly predicted longer NAN time in Omicron patients.

**Conclusion:** This study comprehensively evaluated the similarities and differences in clinical characteristics between SARS-CoV-2 Omicron infection and 2009 H1N1 influenza infection, which is of great significance for a better understanding for these diseases.

## 1. Introduction

The outbreak of COVID-19 in 2019 has created a global pandemic that has infected 664 million patients and killed 6.7 million as of January 22, 2023^[1]^. Because of the strong mutation of COVID-19^[2]^, although we have taken various measures to control the spread of COVID-19, the effect is limited. Alpha variant first appeared, followed by Beta and Delta variant. At present, omicron variant, which was discovered in November 2021, is popular worldwide^[3]^. The data^[4]^ show that the disease and death caused by Omicron variant are significantly lower than the previous ones, but it spreads rapidly (R0 value is close to 10). And it has a high reinfection rate^[5]^ and strong resistance to existing vaccines^[6]^, which poses severe challenges for epidemic prevention and control.

Influenza virus has long been the most common pathogen in respiratory virus infection^[7]^. In the past century, there have been several influenza pandemics^[8]^, one of which was the H1N1 influenza pandemic that began in the spring of 2009 and caused about 284,400 deaths worldwide. [9]The huge death toll from the H1N1 influenza is a wake-up call, while it has many similarities with COVID-19, including similar transmission route, clinical manifestations and transmission range, which provide a reference for dealing with the current global pandemic of the novel coronavirus.

Most previous studies have focused on the difference between wild-type COVID-19 strains and seasonal influenza.^[10,11]^ But COVID-19 is evolving, and the difference between Omicron and seasonal flu is less reported. With the arrival of winter and spring, Omicron and H1N1 influenza may occur at any time. In the current environment where most countries cancel normalized nucleic acid testing, it is of great significance for clinicians to understand the identification of the above two diseases. Besides, most of the previous studies focused on elderly patients^[12,13]^, and the difference between Omicron and H1N1 influenza in young patients is rarely reported. Although young patients are often mild^[14]^, they should also be noticed. Therefore, we analyzed the differences between the patients with H1N1 influenza outbreak in 2009 and the patients with Omicron BA.2 outbreak in 2022 (mainly people under 65 years old), in order to help clinicians better identify H1N1 influenza and Omicron BA.2 infection.

## 2. Methods

### Study subject

From May 2009 to July 2009, a total of 127 patients with H1N1 influenza were diagnosed in Fujian Province. Among them, 126 patients were confirmed to be positive by RT-PCR and Real-time RT-PCR of pharyngeal (nasal) test specimens by Fujian Provincial CDC. One patient was diagnosed by tracking serum antibody titer > 1: 4.

And we also recorded a total of 3265 patients with Omicron diagnosed from March 2022 to May 2022. The second-generation whole genome sequencing was performed by Fujian Provincial Center for Disease Control and Prevention (CDC) for the positive SARS-CoV-2 specimens by fluorescence real-time PCR, and it was confirmed that the novel coronavirus epidemic strain in Quanzhou from March 2022 to May 2022 was Omicron BA.2.

Based on age (The difference is less than 2 years), gender and underlying diseases, we performed a 1 : 2 match between the H1N1 influenza patients and the Omicron patients. Finally, 115 patients with H1N1 and 230 patients with Omicron (referred to as H1N1 group and Omicron group) were included in the statistics.

### Data involvement

Clinical and experimental data were collected from electronic medical records using standardized data collection tables. Serological results included whole blood cells, biochemical tests, and C-reactive protein tests. All results were measured within 24 hours after admission. The time of nucleic acid negative (NAN) was defined the time from the beginning of the patient ‘s course to the first nucleic acid test result being negative.

### Clinical management

In the Omicron group, the asymptomatic and mild patients were quarantined in the mobile cabin hospital (MCH) while patients with moderate or severer symptoms were hospitalized for treatment. All H1N1 influenza patients were treated in hospitals. Both groups of patients were treated according to the guidelines^[15,16]^. Nematavir/Ritonavir antiviral therapy is used for patients who meet the indications. Antibiotic therapy is used for patients considering with bacterial infection.

### Statistical Analysis

For descriptive analysis, we presented data as median [interquartile range (IQR)] for continuous parameters and as frequency (percentage) for categorical variables. Wilcoxon rank sum testing was employed for numerical variables and Fisher’s precision probability test was adopted for categorical variables. Logistic regression was performed to analyze the possible independent risk factors of H1N1 group and Omicron group. And multiple linear regression was used to analyze the influencing factors of NAN time in the two groups. Statistical analysis was performed using SPSS 25.0 data analysis software. A two-tailed P value of less than 0.05 was considered statistically significant.

## 3. Results

### Baseline information

The baseline data of the two groups were shown in TABLE 1. Due to the matching, the median age of both groups was 21[11, 26] years. Among them, 36.52 % were less than 18 years old, 63.48 % were 18-65 years old, and no patients were more than 65 years old. In the gender stratification, men accounted for 56.52 % and women accounted for 43.48 %. And the two groups were not found such as pregnancy, hypertension, diabetes, cardiovascular disease, chronic liver disease, respiratory disease, nervous system disease, metabolic system disease, chronic kidney disease, tumor, etc. Besides, in the Omicron group, the proportion of patients who received 0,1,2, and 3 doses of vaccine was 16.52 %, 6.52 %, 50.87 %, and 26.09 %, respectively.

**TABLE 1.** Baseline information of H1N1 group and Omicron group.

|  | H1N1 group |  | Omicron group |  |
| --- | --- | --- | --- | --- |
|  | N1 | Frequency/IQR | N2 | Frequency/IQR |
| Total(n) | 115 | 100.00% | 230 | 100.00% |
| **Age** |  |  |  |  |
| median age |  | 21[11,26] |  | 21[11,26] |
| < 18 | 42 | 36.52% | 87 | 37.83% |
| 18-65 | 73 | 63.48% | 143 | 62.17% |
| > 65 | 0 | 0.00% | 0 | 0.00% |
| **Sex** |  |  |  |  |
| Male | 65 | 56.52% | 130 | 56.52% |
| Female | 50 | 43.48% | 100 | 43.48% |
| **Underlying diseases** |  |  |  |  |
| Pregnancy | 0 | 0.00% | 0 | 0.00% |
| Hypertension | 0 | 0.00% | 0 | 0.00% |
| Diabetes | 0 | 0.00% | 0 | 0.00% |
| Cardiovascular diseases | 0 | 0.00% | 0 | 0.00% |
| Chronic liver diseases | 0 | 0.00% | 0 | 0.00% |
| Respiratory diseases | 0 | 0.00% | 0 | 0.00% |
| Neurological diseases | 0 | 0.00% | 0 | 0.00% |
| Hematological diseases | 0 | 0.00% | 0 | 0.00% |
| Chronic kidney diseases | 0 | 0.00% | 0 | 0.00% |
| Metabolic diseases | 0 | 0.00% | 0 | 0.00% |
| Tumor | 0 | 0.00% | 0 | 0.00% |
| **Vaccine status (%)** |  |  |  |  |
| No vaccination | - | - | 38 | 16.52% |
| One-dose | - | - | 15 | 6.52% |
| Two-dose | - | - | 117 | 50.87% |
| Three-dose | - | - | 60 | 26.09% |

### Comparison of clinical characteristics

Compared with the Omicron group, the H1N1 group had a higher probability of fever (90.43% VS 34.35%;P<0.001), nasal congestion (12.17% VS 1.74%; P<0.001), sore throat (41.74% VS 21.30%; P<0.001), cough(68.70% VS 36.52%; P<0.001), expectoration(26.09% VS 14.35%; P=0.012), headache (15.65% VS 0.87%; P<0.001), and a lower probability of olfactory loss (0.00% VS 6.96%; P=0.002) and muscle soreness (4.35% VS 14.78%; P=0.004).(TABLE 2)

**TABLE 2.** Comparison of clinical characteristics between the H1N1 and Omicron group.

|  | H1N1 group |  | Omicron group |  | P-value |
| --- | --- | --- | --- | --- | --- |
| total(n) | 115 | 100.00% | 230 | 100.00% |  |
| **clinical features** |  |  |  |  |  |
| Fever n (%) | 104 | 90.43% | 79 | 34.35% | <0.001 |
| Nasal congestion n (%) | 14 | 12.17% | 4 | 1.74% | <0.001 |
| Sore throat n (%) | 48 | 41.74% | 49 | 21.30% | <0.001 |
| Olfactory loss n (%) | 0 | 0.00% | 16 | 6.96% | 0.002 |
| Taste loss n (%) | 0 | 0.00% | 7 | 3.04% | 0.100 |
| Cough n (%) | 79 | 68.70% | 84 | 36.52% | <0.001 |
| Expectoration n (%) | 30 | 26.09% | 33 | 14.35% | 0.012 |
| Fatigue n (%) | 15 | 13.04% | 36 | 15.65% | 0.630 |
| Dyspnea n (%) | 0 | 0.00% | 7 | 3.04% | 0.100 |
| Diarrhea n (%) | 1 | 0.87% | 9 | 3.91% | 0.174 |
| Vomiting n (%) | 1 | 0.87% | 0 | 0.00% | 0.333 |
| Headache n (%) | 18 | 15.65% | 2 | 0.87% | <0.001 |
| Muscle soreness n (%) | 5 | 4.35% | 34 | 14.78% | 0.004 |
| **Laboratory diagnostics** |  |  |  |  |  |
| WBC count (10<sup>9</sup>/L), Median(IQR) | 5.49 | [4.40,6.93] | 4.85 | [3.76,6.45] | 0.008 |
| Lymphocyte count (10<sup>9</sup>/L), Median(IQR) | 1.51 | [1.09,2.02] | 1.58 | [1.09,2.53] | 0.161 |
| Abnormal LDH, n (%) | 21 | 18.26% | 68 | 29.57% | 0.023 |
| CRP (mg/L), Median(IQR) | 4.6 | [2.30,10.45] | 3.23 | [0.51,6.84] | 0.001 |
| **Imaging diagnostics** |  |  |  |  |  |
| pneumonia n (%) | 4 | 3.48% | 16 | 6.96% | 0.229 |
| **Treatment and prognostic indicator** |  |  |  |  |  |
| Antibiotic usage rate, n(%) | 16 | 13.91% | 2 | 0.87% | <0.001 |
| Antiviral usage rate, n(%) | 115 | 100.00% | 2 | 0.87% | <0.001 |
| Nucleic acid negative time (d), Median(IQR) | 4 | [3,5] | 17 | [14,20] | <0.001 |

Among them, fever (90.43%), cough (68.70%) and sore throat (41.74%) were the most common symptoms in H1N1 group. Similarly, the most common symptoms in Omicron group were cough (36.52%), fever (34.35%) and sore throat (21.30%). (Figure 1)

**Figure 1.**
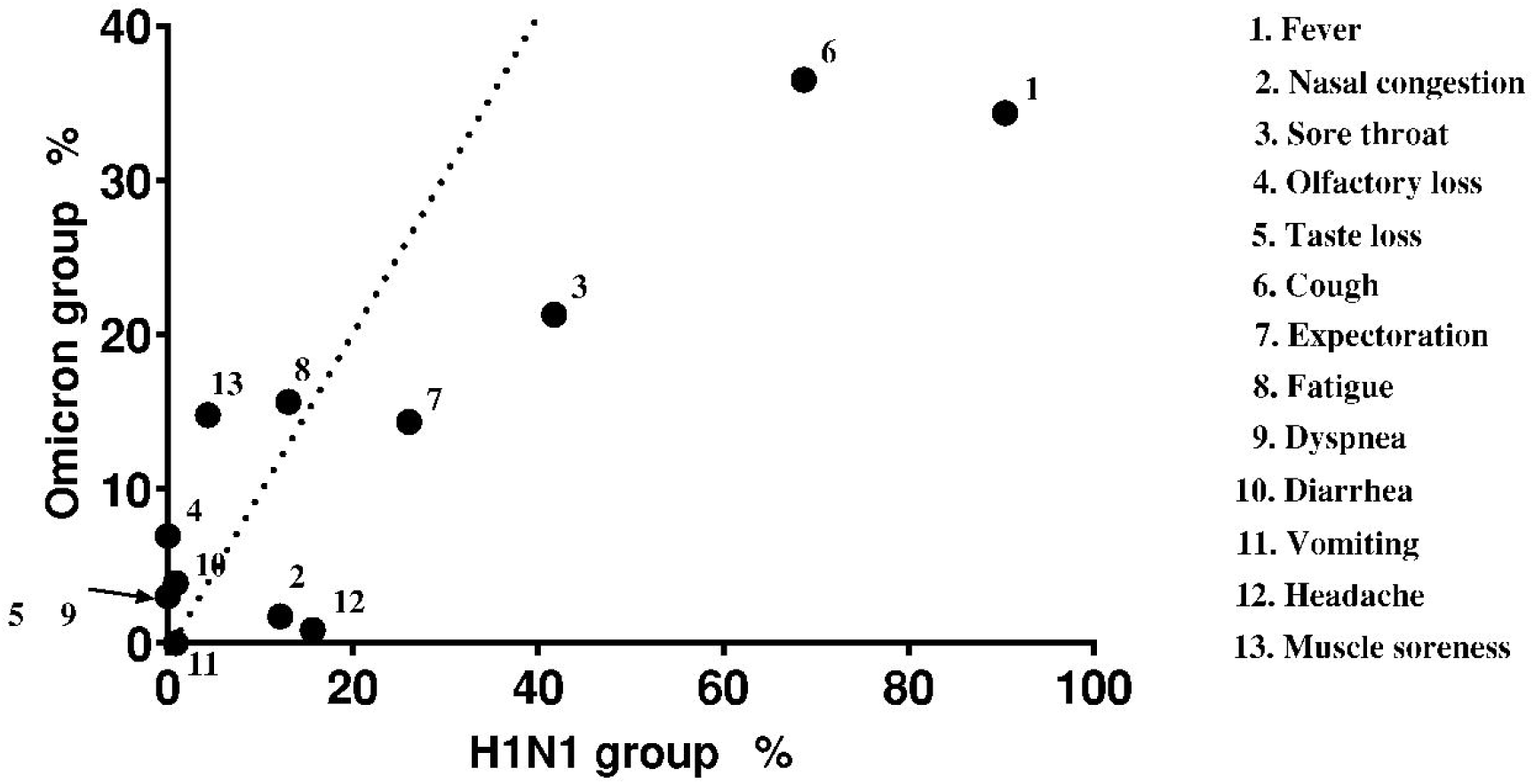
The abscissa of each point represents the frequency (%) of the symptom shown in the H1N1 group, and the ordinate represents the frequency (%) of the symptom shown in the Omicron group. The dotted line indicates that the abscissa and the ordinate are equal here. The arrow indicates that there are completely coincident points here. Fever (90.43% VS 34.35%; P<0.001), cough (68.70% VS 36.52%; P<0.001) and sore throat (41.74% VS 21.30%; P<0.001) were the most common symptoms in both groups, and Omicron group has a lower frequency of symptoms.

The H1N1 group had higher white blood cell count (5.49[4.40,6.93] VS 4.85 [3.76,6.45]; P=0.008) and CRP levels (4.60[2.30,10.45] VS 3.23[0.51,6.84]; P<0.001) (Figure 2), while the LDH abnormal rate (18.26% VS 29.57%; P=0.023) was lower. There was no significant difference in lymphocyte count (1.51 [1.09,2.02] VS 1.58[1.09,2.53]; P=0.161).

**Figure 2.**
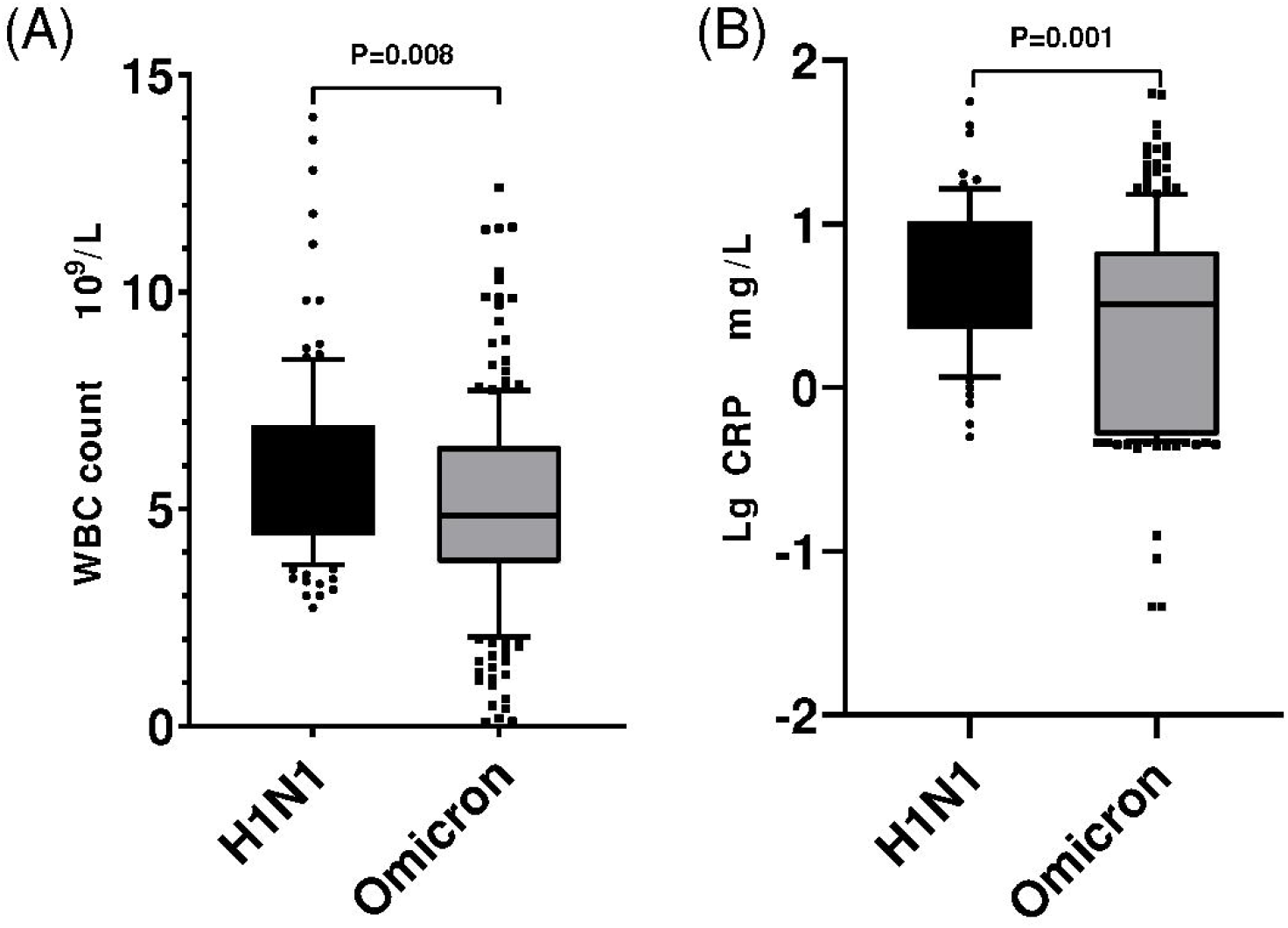
Box plot graphs revealing statistically significant differences in both the white blood cell (WBC) counts (A) and CRP levels (B) between Omicron group and H1N1 group. Most patients with both diseases had normal WBC counts and CRP levels; however, the overall values in H1N1group were higher than those in Omicron (p < 0.05).

In terms of treatment and prognosis, the use of antibiotics (13.91% VS 0.87%; P<0.001) and antiviral drugs (100.00% VS 0.87%; P<0.001) in the H1N1 group was significantly higher than that in the Omicron group. And the time of NAN (4[3,5] VS 17[14,20]; P<0.001) in H1N1 group was significantly lower than that in Omicron group.

### Analysis of risk factors

The indicators with P < 0.05 in the above clinical features, laboratory and imaging diagnostics were analyzed, and the Omicron group was used as a reference. The results showed that the H1N1 group was more prone to fever (OR 19.179, 95%CI [8.82-41.708]; P<0.001), cough (OR 3.541, 95%CI [1.725-7.270]; P=0.001), headache (OR 15.695, 95%CI [2.288-107.679]; P=0.005), elevated white blood cell count (OR 1.190, 95%CI [1.027-1.378]; P=0.020), and less prone to muscle soreness(OR 0.051, 95%CI [0.013-0.200]; P=0.001) and LDH abnormalities (OR 0.393, 95%CI [0.194-0.795]; P=0.009).(TABLE 3)

**TABLE 3.** Analysis of risk factors between H1N1 group and Omicron group

| Factor | B | SE | Wald | P | OR | 95%CI |
| --- | --- | --- | --- | --- | --- | --- |
| Fever | 2.954 | 0.396 | 55.538 | < 0.001 | 19.179 | 8.82-41.708 |
| Nasal congestion | 1.179 | 0.76 | 2.403 | 0.121 | 3.25 | 0.732-14.418 |
| Sore throat | 0.363 | 0.354 | 1.048 | 0.306 | 1.437 | 0.718-2.878 |
| Cough | 1.264 | 0.367 | 11.868 | 0.001 | 3.541 | 1.725-7.270 |
| Expectoration | -0.193 | 0.459 | 0.176 | 0.675 | 0.825 | 0.335-2.030 |
| Headache | 2.753 | 0.983 | 7.852 | 0.005 | 15.695 | 2.288-107.679 |
| Muscle soreness | -2.971 | 0.695 | 18.281 | < 0.001 | 0.051 | 0.013-0.200 |
| WBC count | 0.174 | 0.075 | 5.39 | 0.02 | 1.19 | 1.027-1.378 |
| Abnormal LDH | -0.934 | 0.359 | 6.759 | 0.009 | 0.393 | 0.194-0.795 |
| CRP | 0.009 | 0.018 | 0.287 | 0.592 | 1.009 | 0.975-1.045 |

Multivariate logistic regression analysis was performed with Omicron group as reference. Missing data are processed by multiple interpolation.

### Analysis of influencing factors of NAN time

The results showed that for H1N1 patients, elevated WBC count significantly predicted longer NAN time (B 0.217, 95%CI [0.028,0.406]; P=0.025), while fatigue was associated with shorter one (B -1.589, 95%CI [-2.646, -0.532]; P=0.004). For Omicron patients, fever significantly predicted longer NAN time (B 1.529, 95%CI [0.149,2.909]; P=0.030). (TABLE 4)

**TABLE 4.** Analysis of influencing factors for the time for nucleic acid negativization (NAN)

| Variables | H1N1 |  |  |  | Omicron |  |  |  |
| --- | --- | --- | --- | --- | --- | --- | --- | --- |
|  | Univariate |  | Multivariate |  | Univariate |  | Multivariate |  |
|  | B | P value | B (95%CI) | P value | B | P value | B (95%CI) | P value |
| Fever | 0.625 | 0.353 |  |  | 1.583 | 0.023 | 1.529<br>(0.149,2.909) | 0.030 |
| Nasal congestion | -0.133 | 0.826 |  |  | 0.816 | 0.749 |  |  |
| Sore throat | -0.009 | 0.982 |  |  | 0.066 | 0.935 |  |  |
| Olfactory loss | - | - |  |  | 1.064 | 0.416 |  |  |
| taste loss | - | - |  |  | 1.233 | 0.525 |  |  |
| Cough | -0.038 | 0.929 |  |  | 0.514 | 0.458 |  |  |
| Expectoration | 0.08 | 0.859 |  |  | 0.379 | 0.690 |  |  |
| Fatigue | -1.503 | 0.009 | -1.589<br>(-2.646, -0.532) | 0.004 | 1.346 | 0.141 | 1.077<br>(-0.724,2.878) | 0.240 |
| Dyspnea | - | - |  |  | 0.054 | 0.978 |  |  |
| Diarrhea | 0.531 | 0.803 |  |  | 0.054 | 0.975 |  |  |
| Vomiting | -2.496 | 0.241 |  |  | - | - |  |  |
| Headache | -1.089 | 0.044 | -0.986<br>(-1.978, -0.006) | 0.051 | 1.566 | 0.663 |  |  |
| Muscle soreness | 0.968 | 0.318 |  |  | 1.166 | 0.214 |  |  |
| WBC count | 0.325 | 0.001 | 0.217<br>(0.028,0.406) | 0.025 | -0.063 | 0.673 |  |  |
| Lymphocyte count | 0.074 | 0.657 |  |  | -0.093 | 0.665 |  |  |
| Abnormal LDH | 0.639 | 0.170 | 0.182<br>(-0.666,1.029) | 0.672 | -0.015 | 0.983 |  |  |
| CRP | 0.049 | 0.026 | 0.03<br>(-0.011,0.071) | 0.152 | 0.040 | 0.297 |  |  |
| Pneumonia<br>(Imaging) | 2.099 | 0.05 | 1.473<br>(-0.525,3.470) | 0.147 | -0.347 | 0.791 |  |  |
| Antibiotic<br>usage rate | 1.809 | 0.001 | 1.076<br>(-0.100,2.252) | 0.072 | 5.601 | 0.118 | 6.301<br>(-0.666,13.267) | 0.076 |
| Antiviral<br>usage rate | - | - |  |  | -2.469 | 0.491 |  |  |

The factors affecting NAN time in the H1N1 group and Omicron group were analyzed by multiple linear regression. Indices of statistical differences( $P < 0.200$ )univariate analysis were included in the multivariate regression analysis. “-” indicates that too many or too few values to perform single factor regression analysis. Missing data are processed by multiple interpolation.

## 4. Discussion

The novel coronavirus, which began in 2019, is constantly spreading and mutating, having a huge impact on the disease spectrum and bringing huge burden to people worldwide^[17]^. In winter and spring, seasonal influenza virus infection is also very common. It is a respiratory virus different from SARS-CoV-2, but the clinical characteristics of patients infected with it are similar, which brings lots of difficulties to the diagnosis and treatment. Besides, there is a chance of co-infection^[18]^. Therefore, we intend to learn from the previous experience in dealing with the 2009 H1N1 influenza pandemic^[19]^, and compare it with the Omicron epidemic in Quanzhou^[20]^, which may better enable us to identify the two diseases.

Due to the younger age and fewer underlying diseases in the H1N1 group, the matched Omicron group also had such characteristics. The median age of the Omicron group was 21 [11,26] years, which was significantly lower than the median age of 36 [25,48] years in our previous study^[20]^. The younger age of the H1N1 group may be related to the characteristics of the virus^[21]^ (all 127 H1N1 influenza patients were younger than 65 years old). Since the outbreak of H1N1 influenza in China was in the early stage of epidemic and most patients had not been vaccinated, the effect of vaccination in these two groups was not studied.

Our study found that fever (90.43% VS 34.35%; P<0.001), cough (68.70% VS 36.52%; P<0.001) and sore throat (41.74% VS 21.30%; P<0.001) were the most common symptoms in both groups, consistent with previous studies^[22,23]^ that respiratory symptoms were the main symptoms. And the frequency of symptoms in the H1N1 group was higher than that in the Omicron group, which was also consistent with the previous study that the virulence of Omicron decreased and showed more asymptomatic^[24]^. The probability of muscle soreness (14.78 % VS 4.35 %) was higher after Omicron infection, suggesting that Omicron was more likely to invade muscles. This may be related to the expression of ACE-2 receptor in skeletal muscle cell and other cells in muscle such as satellite cells, white blood cells, fibroblasts and endothelial cells. In addition to immune-mediated muscle damage, Omicron may also directly invade skeletal muscle and cause muscle damage^[25]^.

Omicron group had symptoms of olfactory loss and taste loss, which were major features different from H1N1 group. The possible mechanism is that Omicron adheres to the motor cilia with the help of ACE-2 receptor, breaks through the periciliary layer (PCL)^[26]^, infiltrates the olfactory epithelial tissue and induces local inflammatory response^[27]^, eventually causing microvascular and axonal changes^[28]^ and affecting the expression of olfactory related genes^[29]^. And there are studies shows that in nasal tissue, the body ‘s response to SARS-CoV-2 is more extensive than that to influenza virus, including the maturation and activation of immune cells in both innate immunity and adaptive immunity^[30]^, which leads to a stronger immune response in the nasal mucosa and causes greater damage, making for a decline in olfactory function. Omicron can also enter specific epithelial taste cells through ACE-2 receptor and destroy normal taste function^[31]^. Or by affecting the oral symbiotic flora to change the immune status and induce cytokine storms, ultimately damaging the taste nerve and destroying the taste function^[32]^. Although there is no effective treatment for sensory loss^[27]^, most patients (>95%) can recover completely or nearly completely within half a year after the acute phase^[33]^.

Our study showed that H1N1 group had higher white blood cell count and CRP levels, which may suggest that the inflammatory response caused by H1N1 influenza is more severe. Different from previous studies^[11]^, the white blood cell count, lymphocyte count and CRP level in the Omicron group were within the normal range, which may be related to the younger age of the included population. The immunity of elderly patients is weak. With the increase of age, the diversity of T cells, which plays an important role in eliminating virus^[34]^, gradually decreases. And after 40 years old, it shows a trend of sharp decline^[35]^. Besides, there was a study showed that in Omicron patients, young people had higher levels of neutralizing antibodies^[36]^. The above differences may allow the virus in young patients to be cleared faster without significantly affecting the immune system, so that the lymphocyte count is still within the normal range, and the clinical symptoms of young patients are relatively mild compared to the elderly^[37]^. Omicron group had higher probability of LDH abnormality (29.57% VS 18.26%), which may be related to the wide distribution of ACE-2 receptor in body tissues^[38]^. Omicron may cause tissue damage through this receptor, leading to muscle soreness and abnormal LDH, but the specific mechanism is not clear.

In our study, whether H1N1 patients or Omicron patients, we found that the incidence of pneumonia in young patients was lower (3.48% VS 6.96%, P=0.229), and the symptoms were mainly upper respiratory tract infection. On the one hand, this may be related to the age characteristics of the patient, the younger the patient, the lighter the condition^[37]^; on the other hand, it may be related to the relatively strong nasal and weakened pulmonary tropism of Omicron^[26]^. In addition, since H1N1 is one of the influenza viruses, there was a specific antiviral drug oseltamivir when it broke out. The high utilization rate of antiviral drugs (100.00 % VS 0.87 % ; p < 0.001) may lead to low incidence of pneumonia. Besides, the use of antibiotics in the H1N1 group was higher than that in the Omicron group, indicating a higher probability of bacteria co-infection, which was consistent with the findings of previous studies^[39]^.

By Logistic regression analysis, we found if the patient had fever, cough, headache, or 1 unit (10^9/L) increasement of white blood cell count, the probability of H1N1 was 18.689, 3.853, 16.649, 1.228 times higher than that of Omicron. This was obviously different from previous studies^[40,41]^ in which there were more symptoms such as fever and cough after COVID-19 wild strain infection. H1N1 was less likely to cause muscle soreness and LDH abnormalities (the probability was 0.102 and 0.373 times that of Omicron, respectively). The specific reason may be related to the damage of Omicron to many body tissues including muscle tissue because of the widely distributed ACE-2 receptor^[25,38]^.

In our study, we found that the NAN time of Omicron group was significantly higher than that of H1N1 group (17 [14,20] VS 4 [3,5] days, P < 0.001). Negative nucleic acid means that the patient is no longer the source of infection. And a longer NAN time shows that Omicron has a longer infectious time and spreads faster than H1N1 (R0 value :10 vs 2.75)^[42,43]^. The reason may be related to the specific antibody production and the use of antiviral drugs. Due to the long-term seasonal influenza epidemic in the population, some people have cross-antibody against H1N1^[44]^. Moreover, specific anti-H1N1 virus drugs can quickly inhibit the virus, so that H1N1 virus can be quickly cleared in vivo. However, Omicron often has immune escape due to its strong mutation ability^[45]^, and the ability of neutralizing antibodies produced by the body to Omicron strain is significantly reduced^[36]^. Besides, the antiviral effect of Omicron is limited, which leads to a significant prolongation of virus clearance time. During the H1N1 epidemic, based on the use of the specific antiviral drug oseltamivir^[19]^, the symptoms of most patients were relieved quickly and the time of NAN was greatly shortened. Nevertheless, during the Omicron epidemic, since the Paxlovid had not been popularized in China, the use rate of antiviral drugs remained low^[20]^, which made the nucleic acid turn negative for a long time. In addition, this study analyzed the risk factors of NAN time in the two groups of patients and found that the antibiotic use and the increased white blood cell count suggested longer NAN time, which maybe due to the bacteria co-infection making the condition more complicated^[46]^. For Omicron patients, fever is positively correlated with the time of NAN. All these suggest that if H1N1 patients have elevated hemogram and Omicron patients have fever, we should be prepared for a longer course of disease and early use of antiviral drugs may be helpful for shortening the time of NAN^[47,48]^.

Our study also has some limitations: (1) All the included people are less than 65 years old, which cannot reflect the situation of high-risk people who has advanced age and many basic diseases. (2) Because this study was a retrospective study, some clinical data were missing, such as fever duration, antibodies, etc., making it hard to to better compare the immune status of all patients. (3) Due to the large biological differences between H1N1 and Omicron, the influencing factors of NAN time are complicated and there must be bias in the analysis.

## Data Availability

The original contributions presented in the study are included in the article/Supplementary Material. Further inquiries can be directed to the corresponding author.

## Ethics Statement

The studies involving human participants were reviewed and approved by Ethics Committee of Fujian Provincial Hospital (ethics number: K2019-12-032) and the Ethics Committee of Quanzhou First Hospital (ethics number: No202212). Written informed consent to participate in this study was provided by the participants’ legal guardian/next of kin. Written informed consent was obtained from the individual(s), and minor(s)’ legal guardian/next of kin, for the publication of any potentially identifiable images or data included in this article.

## Author contributions

HRL, HL were responsible for funding acquisition. HRL, YWX were responsible for design conception. JBF were responsible for formal analysis. WZ, YSW, JBF performed data curation. HL, YSC were responsible for resources. BSX, NLX, ML, XPZ verified the underlying data. YSW, WZ prepared the original draft. All authors contributed to manuscript reviewing and editing. All authors had access to all data in the study and the corresponding author had final responsibility for the decision to submit the paper for publication

## Acknowledgments

This study was supported by grants from the National Science and Technology Major Special Project (No.2017Z 10103004), Natural Science Foundation of Fujian Province (No.2019J01178), and Fujian Provincial Hospital “Creating double High” Flint Fund project (No.2019HSJJ11).

